# Effect of Heat inactivation on Real-Time Reverse Transcription PCR of the SARS-COV-2 Detection

**DOI:** 10.1101/2020.05.19.20101469

**Authors:** Yanxia Liu, Zhengan Cao, Mei Chen, Yan Zhong, Yuhao Luo, Gang Shi, Hongxia Xiang, Jianping Luo, Hongwei Zhou

**Affiliations:** Microbiome Medicine Center, Division of Laboratory Medicine, Zhujiang Hospital, Southern Medical University, Guangzhou, Guangdong, China; Zhuzhou center for Disease Control and Prevention, Zhuzhou, Hunan, China

**Keywords:** SARS-CoV-2, Heat Inactivation, rRT-PCR, Comparison

## Abstract

Real-time reverse transcription PCR (rRT-PCR) is commonly used to diagnose SARS-CoV-2 infection. Heat inactivation prior to nucleic acid isolation may allow safe testing, while the effects of heat inactivation on SARS-CoV-2 rRT-PCR detection result need to be determined. 14 positive nasopharyngeal swab specimens were inactivated at 56°C for 30min, 56°C for 60min, 60°C for 30min, 60°C for 75min, and 100°C for 10min, then were detected by rRT-PCR. All 14 heat treated samples remained positive. Another 2 positive nasopharyngeal swab specimens were inactivated at 100°C for 10min, 100°C for 30min, and 100°C for 60min, after which the samples were isolated and detected by rRT-PCR. The range of threshold cycle (Ct) values observed when detecting ORF1a/b was 27.228-34.011 in heat-treated samples, while 25.281-34.861 in unheated samples, and the range of threshold cycle (Ct) values observed at the time of detecting N was 25.777-33.351 in heat-treated samples, while 24.1615-35.433 in unheated samples, on basis of which it showed no statistical difference otherwise a good correlation of Ct values between the heat-inactivated samples and the untreated samples. However, the 2 samples inactivated at 100°C 30min, 100°C 60min turned into negative. Heat inactivation at 56°C for 30min, 56°C for 60min, 60°C for 30min, 60°C for 75min, and 100°C for 10min shall not affect the detection results of Real-Time Reverse Transcription PCR of the SARS-COV2. Furthermore, it is recommended to inactive nasopharyngeal swab specimens 10min at 100°C before RNA extraction in consideration of efficiency and reliable results.

## 1 INTRODUCTION

The rapid spread of severe acute respiratory syndrome coronavirus 2 (SARS-CoV-2) throughout the world has become a major challenge for global public health. Globally, as of 2:00am CEST, 7 Apr 2020, there have been 1,279,722 confirmed cases of COVID-19, including 72,616 deaths, reported to WHO^1^.

SARS-CoV-2 has been deemed to be transmitted from human to human through direct contact (cough, sneeze, and droplet inhalation), or indirect contact (contact with oral, nasal, and eye mucous membranes). Studies have suggested that SARS-CoV-2 may spread through aerosols formed during medical procedures^2-8^.

Early detection has a significant impact on the prevention and control of SARS-CoV-2, by real-time reverse transcription PCR (rRT-PCR) assays in nasopharyngeal swab specimens from infected patients, which would put the laboratory workers at great risk for SARS-CoV-2 infection due to exposure to respiratory, blood, stool specimens and other body fluid specimens. It has given rise participants of virus detection under tremendous psychological pressure as well as huge economic and human cost of protective measurements. Whereas the above, we have been attempting to mitigate the situation through our study.

It has been shown that the coronavirus titers were highly effective reduced to the detection limit at 56°C for 30 min for SARS-CoV and MERS-CoV, and 60°C was also the sensitive temperature^9-11^.

Notwithstanding the foregoing, different protein concentrations may have protective effects and may affect the inactivation efficiency, due to which, it is well-advised to prolong the heating time or increase the temperature to completely inactivate viruses^12-15^. However, because viral RNA would easily be degraded by ribonucleases, which affects the detection result of rRT-PCR of virus.

Systematic research on the heat inactivation efficacies of rRT-PCR remains to be studied and confirmed.

we evaluated the efficacy of SARS-CoV-2 heat inactivation in nasopharyngeal swab specimens in our study, where we performed different commonly used heat condition to inactive the SARS-CoV-2, i.e, 56°C for 30min, 56°C for 60min, 60°C for 30min, 60°C for 75min, 100°C for 10min, 100°C for 30min and 100°C for 60min. And then we evaluated and compared the ct values of heat treated and unheated samples in nasopharyngeal swab specimens.

## 2 Materials and methods

### 2.1 Specimen collection and testing

Nasopharyngeal swab specimens were obtained according to CDC guidelines and collected in a separate sterile tube containing 1 ml of viral transport medium (including guanidine isothiocyanate, sodium dodecylsarcosine, and dithiothreitol).

This study was approved by the ethics commission of Zhujiang Hospital.

The 14 samples were submitted to three temperatures with different durations. Which included: unheated, 56°C for 30min, 56°C for 60min, 60°C for 30min, 60°C for 75min, and 100°C for 10min. While the other 2 samples were submitted to three conditions of 100°C for 10min, 100°C for 30min and 100°C for 60min.

RNA was extracted and tested by real-time RT-PCR using the kit (bioperfectus technologies) according to the manufacturer’s instructions. Tests were performed in biosafety level 2 facilities at the Zhuzhou Center for Disease Control and Prevention.

The sample would be considered to be laboratory-confirmed, if two targets (open reading frame 1a or 1b, nucleocapsid protein) are tested positive by specific real-time RT-PCR. And a cycle threshold value (Ct-value) less than 36 shall be defined as positive, with a Ct-value of 40 or more as negative.

### 2.2 Statistical Analysis

Quantitative results were expressed as mean±SD, and groups were compared by t-test. P values <0.05 was considered statistically significant.

Graphpad Prism software (Version8.0) was used for statistical analysis. The Spearman correlation coefficient was used to analyze the correlation between CT values of different treated samples, and the Bland-Altman plot was used to analyze the consistency of CT values.

## 3 Results

### 3.1 rRT-PCR results of inactivated and non-inactivated samples at different temperatures and durations

The effects of heat treatment at different temperatures and durations on the SARS-CoV-2 rRT-PCR Ct-value were evaluated. As shown in table 1, The Ct values of ORF1a/b from 14 nasopharyngeal swab samples were 25.63-34.86, and the Ct values of the heat inactivated samples were similar to the non-inactivated samples. The average Ct values difference between inactivated samples at 56°C for 30min and non-inactivated samples is −0.7175 to 1.947, 0.102 to 2.806 between inactivated samples at 56°C for 60min and non-inactivated samples, 0.0145 to 2.0435 between inactivated samples at 60°C for 30min and non-inactivated samples, −0.3735 to 2.489, between inactivated samples at 60°C for 60min and non-inactivated samples, −1.321 to 2.254 between inactivated samples at 100°C for 10min and non-inactivated samples. On basis of which, only the Ct values of sample 2 and 4 were less than the non-inactivated ones, while others were more than the non-inactivated ones, i.e, all Ct values of samples at 56°C for 60mins and 60°C for 30mins were more than the non-inactivated ones. Notwithstanding the foregoing, all heat inactivated samples were tested positive.

**Table 1.**
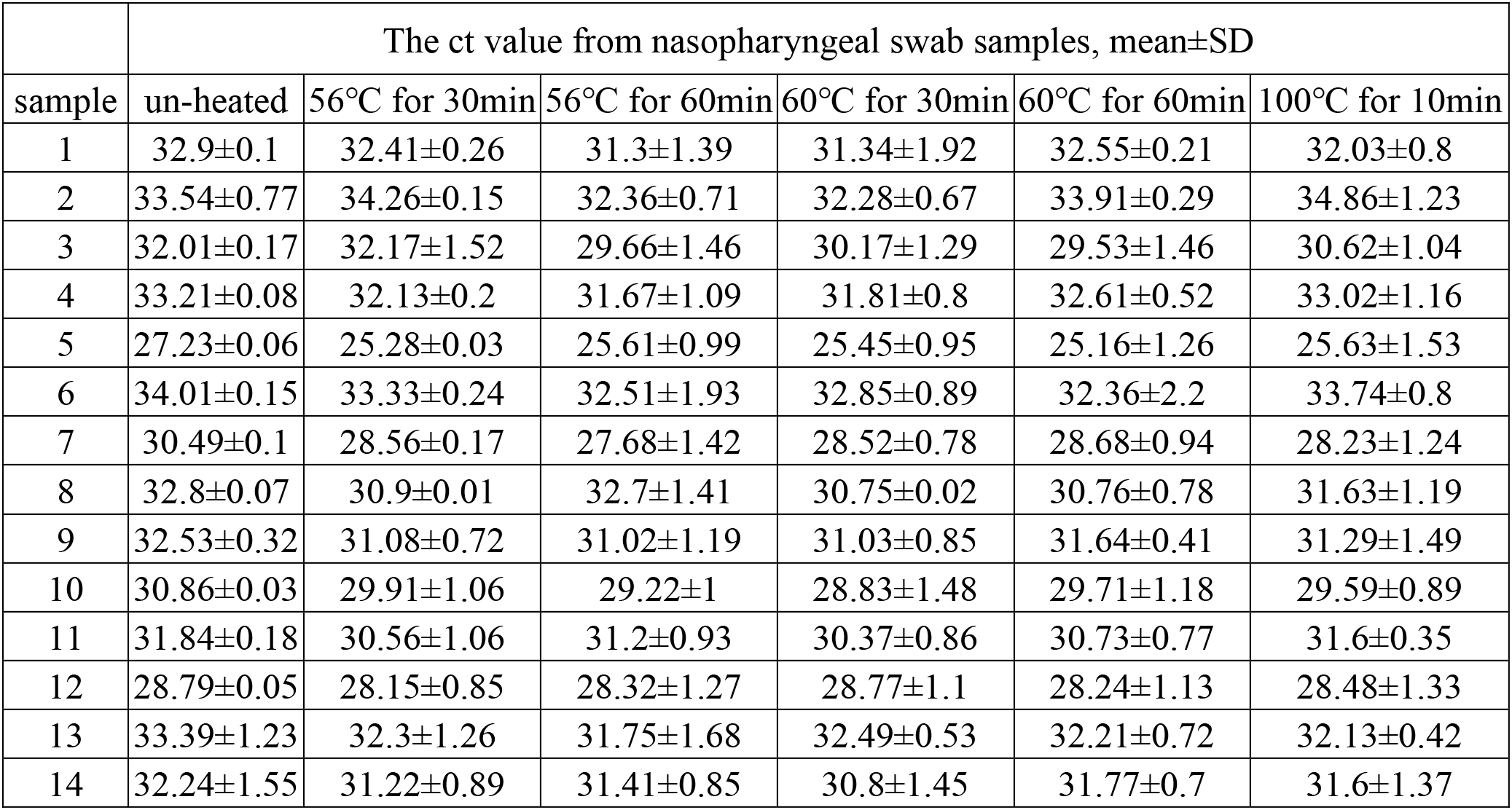
Cycle threshold (Ct) values for ORF1a/b measured with nasopharyngeal swab samples following heat treatments at different temperatures and durations

As shown in table 2, The Ct values of N of 14 nasopharyngeal swab samples were 24.46-35.43, and the Ct values of the heat-inactivated samples were approximate to the non-inactivated samples. The average Ct values difference between the inactivated samples at 56°C for 30min and the non-inactivated samples is −0.359 to 2.019, −0.3385 to 2.088 between the inactivated samples at 56°C for 60min and the non-inactivated samples, −0.646 to 1.7505 between the inactivated samples at 60°C for 30min and the non-inactivated samples, −0.1625 to 1.921 between the inactivated samples at 60°C for 60min and the non-inactivated samples, −2.082 to 1.418 between the inactivated samples at 100°C for 10min and non-inactivated samples.

**Table 2.** Cycle threshold (Ct) values for N measured with nasopharyngeal swab samples following heat treatments at different temperatures and drations

|  | The ct value from nasopharyngeal swab samples, mean±SD |  |  |  |  |  |
| --- | --- | --- | --- | --- | --- | --- |
| sample | un-heated | 56°C for 30min | 56°C for 60min | 60°C for 30min | 60°C for 60min | 100°C for 10min |
| 1 | 31.88±0.17 | 31.44±0.12 | 30.98±1.43 | 30.98±1.51 | 31.87±0.89 | 31.74±1.01 |
| 2 | 33.35±0.43 | 32.65±0.34 | 31.63±1.54 | 31.89±0.67 | 32.84±0.89 | 35.43±1.48 |
| 3 | 30.82±0.16 | 31.18±1.22 | 29.19±1.83 | 29.89±1.4 | 29.67±1.4 | 30.28±1.17 |
| 4 | 32.16±0.25 | 31.27±0.1 | 31.65±0.7 | 30.81±0.45 | 32.29±0.47 | 33.13±0.55 |
| 5 | 25.78±0.03 | 24.16±0.03 | 24.85±1.03 | 24.58±1.01 | 24.46±1.28 | 25.2±1.18 |
| 6 | 32.67±0.2 | 32.48±0.26 | 32.29±1.5 | 32.76±0.32 | 32.08±1.51 | 33.56±0.97 |
| 7 | 29.41±0.01 | 27.93±0.03 | 27.32±1.19 | 28.13±0.9 | 28.68±1.17 | 27.99±1.11 |
| 8 | 32.29±0.01 | 30.27±0.08 | 31.69±1.73 | 30.54±0.58 | 30.37±1 | 31.64±1.35 |
| 9 | 31.36±0.47 | 30.31±0.94 | 30.28±1.86 | 30.8±1.15 | 31.04±1.03 | 30.55±1.3 |
| 10 | 29.95±0.05 | 29.66±1 | 28.84±1.22 | 29.06±1.63 | 30.11±0.53 | 29.55±1.36 |
| 11 | 30.97±0.11 | 30.43±1.08 | 30.66±1.03 | 30.12±1.05 | 30.28±1.15 | 31.24±0.54 |
| 12 | 27.52±0.05 | 27.55±0.93 | 27.86±1.36 | 28.17±1.17 | 27.52±1.25 | 27.97±1.53 |
| 13 | 32.28±1.19 | 31.6±1.21 | 31.32±1.4 | 31.95±0.79 | 31.99±1.12 | 31.86±1.02 |
| 14 | 31.55±1.03 | 31.21±1.01 | 31.05±0.92 | 30.59±1.22 | 31±0.74 | 31.63±1.26 |

As shown from the tables above, there was no statistical difference in Ct values between the heat-inactivated and non-inactivated samples, though most Ct values tended to be higher than untreated samples after heat treatments.

### 3.2 Correlation Analysis of qPCR Results of the Inactivated and non-Inactivated Samples

The ct values of the heat treatments samples (at 56°C for 30 min, 56°C for 60 min, 60°C for 30 min, 60°C for 75 min, 100°C for 10 min) were significantly related to the non-inactivated samples. Spearman correlation coefficient indicated that r = P <0.001 (Fig.1).

**Fig 1.**
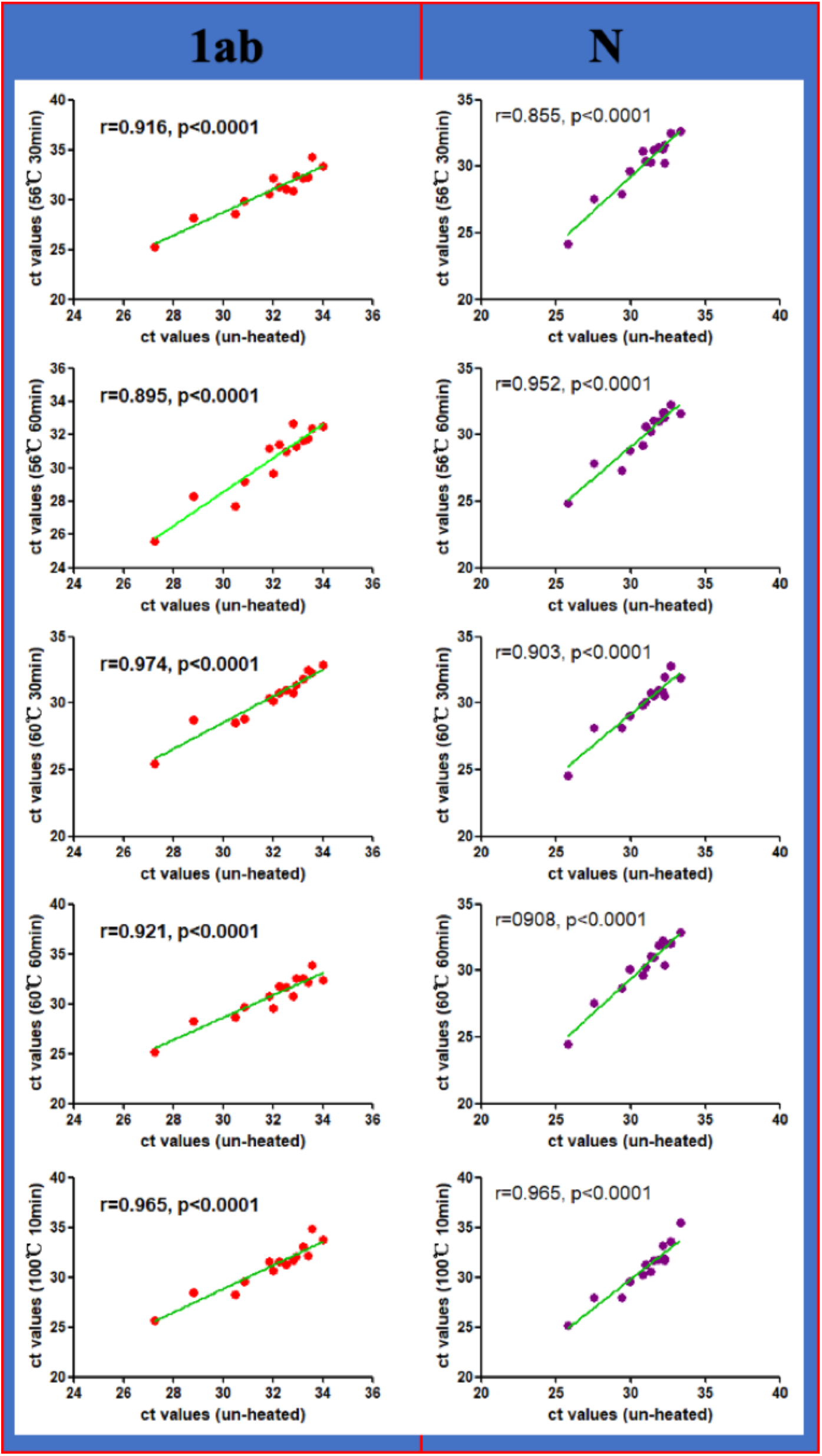
Spearman correlation analysis of The Ct values for ORF1a/b and N between Inactivated and non-Inactivated Samples

### 3.3 rRT-PCR results of inactivated at 100°C

Since there is no statistical difference between non-inactivated and the above activated conditions, two more samples were performed to compare the results between the non-inactivated samples and the activated ones at 100°C for 10 min, 30 min, and 60 min. As shown in table 3, the virus could not be detected in the event of heat treatment at 100°C for 30 mins, and 60 minutes, even if there is no statistical difference between the activated ones at 100°C for 10 mins and the non-inactivated samples.

**Table 3.** Cycle threshold (Ct) values for ORF1a/b and N measured with nasopharyngeal swab samples following heat treatments at 100°C

| gene | sample | The ct value from nasopharyngeal swab samples, mean±SD |  |  |  |
| --- | --- | --- | --- | --- | --- |
|  |  | un-heated | 100°C for 10min | 100°C for 30min | 100°C for 60min |
| ORF1a/b | 15 | 29.50±0.04 | 29.29±0.49 | un-detected | un-detected |
| N |  | 32.03±0.43 | 33.24±0.66 | un-detected | un-detected |
| ORF1a/b | 16 | 31.82±0.16 | 31.94±0.72 | un-detected | un-detected |
| N |  | 32.06±0.45 | 31.97±0.61 | un-detected | un-detected |

## 4 Discussion

SARS-CoV-2 has been emerged in late 2019 in China and now is in the list of Public Health Emergency of International Concern (PHEIC). Respiratory specimens have been used to diagnosis SARS-CoV-2 infection by rRT-PCR, and are regarded as the main detection method and gold standard. During sample testing, it has been strongly recommended to use barrier-protection equipment, including but not limit to protective glasses, face shields, gloves, hats, face shields and protective clothing, resulting in increasing the inconvenience and workload of the experiment.

It has been shown that in the SARS-CoV-2 culture supernatant, the SARS-CoV-2 titers could be reduced at 56°C for 30min and 60°C for 60min, but still infectious. Only heating at 92°C for 15min was able to inactivate it totally^16^. However, the thermostability of SARS-CoV-2 RNA in nasopharyngeal swab specimens is still unknown. The incomplete inactivation of viruses may bring laboratory biosafety risks. Nevertheless, we have been trying to demonstrate that it is possible to ensure the test integrity by applying heat inactivation at several conditions.

Previous studies have found that inactivation at 56°C for 30 min could result in a decreased detectable amount and increased Ct values. In particular, diluted weak positive samples may become false negatives in SARS-CoV-2 RT-PCR detection^17^. However, our study has shown that the virus loads measured in the heat-inactivated samples were consistent with those in the unheat-inactivated ones, with sufficient amounts of RNA preserved for downstream rRT-PCR testing, which concurs with the conclusions of Wilber Sabiiti et al. and Boris Pastori et al^16-18^. The RNA preservation may be due to the preservation solution, which contains guanidine isothiocyanate, sodium dodecylsarcosine, and dithiothreitol. This suggested that the presence of the preservation solution can effectively protect the integrity of the viral nucleic acid, thereby increasing the proportion of detectable nucleic acids. And It may also be that we used the original samples rather than the diluted samples.

Although our study showed that heating at 100°C for 10min was consistent with un-heated ones. Heating at 100°C for 30min and 60min would result in a false negative, which is consistent to that heating at 92°C for 15min, the SARS-CoV-2 RNA copies in the culture supernatant dropped significantly^16^. In fact, studies have suggested that the sample cells shall die and rupture in the event of relatively high temperature and long durations, leading to the release of a large number of cell nucleases, and then a large amount of RNA degradation, which may contribute to false negatives in nucleic acid detection. We hypothesize that heating at 100°C for 30 min or 60 min lyse a large number of cells, and expose RNA to RNases present in the samples, and the duration is also a key factor.

Our research has some limitations. First, only nasopharyngeal swab samples were used for testing; which may make our results not applicable to those obtained using sputum, fecal specimen or blood samples. Second, we tested a relatively small number of samples under limit test condition, affecting the representative significance. Third, the thermostability of SARS-CoV-2 RNA in nasopharyngeal swab specimens at 100 °C for 10 min need to be determined. In the light of which, our findings need to be further verified by other large-scale researches and virus infectivity experiments using cell culture.

## 5 Conclusion

Our finding has showed that the heat inactivation could ensure the test integrity to perform rRT-PCR testing of SARS-CoV-2 samples. Moreover, heating at 100°C for 10 minutes is recommend as the heat inactivation condition for the sake of efficiency. And this study should help laboratory workers to choose the best suited inactivation protocol for them in order to avoid the risk of exposure. Furthermore, it could save manpower and financial resources, improve work efficiency, and reduce the risk of infection. Especially in immature areas, it is conducive to carry out the virus detection.

## Data Availability

all data referred to in the manuscript is availability.

## ACKNOWLEDGMENTS

This work was supported by research grants from the National Natural Science Foundation of China (81925026, and 82041007).

## References

1. WHO. Novel coronavirus – WHO COVID-19 dashboard. https://covid19.who.int. Accessed May 24, 2020.

2. Li Q, Guan X, Wu P, Wang X, Zhou L, Tong Y, Ren R, Leung KSM, Lau EHY, Wong JY et al. Early Transmission Dynamics in Wuhan, China, of Novel Coronavirus-Infected Pneumonia. N Engl J Med. 2020;382(13):1199–1207.

3. Carlos WG, Dela Cruz CS, Cao B, Pasnick S, Jamil S. Novel Wuhan (2019-nCoV) Coronavirus. Am J Respir Crit Care Med. 2020;201(4):P7-p8.

4. Chan JF, Yuan S, Kok KH, To KK, Chu H, Yang J, Xing F, Liu J, Yip CC, Poon RW et al. A familial cluster of pneumonia associated with the 2019 novel coronavirus indicating person-to-person transmission: a study of a family cluster. Lancet. 2020;395(10223):514–523.

5. Lai CC, Shih TP, Ko WC, Tang HJ, Hsueh PR. Severe acute respiratory syndrome coronavirus 2 (SARS-CoV-2) and coronavirus disease-2019 (COVID-19): The epidemic and the challenges. Int J Antimicrob Agents. 2020;55(3):105924.

6. Luo C, Yao L, Zhang L, Yao M, Chen X, Wang Q, Shen H. Possible Transmission of Severe Acute Respiratory Syndrome Coronavirus 2 (SARS-CoV-2) in a Public Bath Center in Huai’an, Jiangsu Province, China. JAMA Network Open. 2020;3(3):e204583-e204583.

7. Singhal T. A Review of Coronavirus Disease-2019 (COVID-19). Indian J Pediatr. 2020;87(4):281–286.

8. Wang D, Hu B, Hu C, et al. Clinical characteristics of 138 hospitalized patients with 2019 novel coronavirus-infected pneumonia in Wuhan, China. JAMA. 2020. https://doi.org/10.1001/jama.2020.1585

9. Leclercq I, Batejat C, Burguiere AM, Manuguerra JC. Heat inactivation of the Middle East respiratory syndrome coronavirus. Influenza Other Respir Viruses. 2014;8(5):585–586.

10. Darnell ME, Taylor DR. Evaluation of inactivation methods for severe acute respiratory syndrome coronavirus in noncellular blood products. Transfusion. 2006;46(10):1770–1777.

11. Rabenau HF, Cinatl J, Morgenstern B, Bauer G, Preiser W, Doerr HW. Stability and inactivation of SARS coronavirus. Med Microbiol Immunol. 2005;194(1-2): 1-6.

12. Darnell ME, Subbarao K, Feinstone SM, et al. Inactivation of the coronavirus that induces severe acute respiratory syndrome, SARS-CoV. Journal of Virological Methods. 2004; 121(1)185-91.

13. Kariwa H, Fujii N, Takashima I. Inactivation of SARS coronavirus by means of povidone-iodine, physical conditions and chemical reagents. Dermatology. 2006;212 Suppl 1(Suppl 1):119–123. doi:10.1159/000089211

14. Yunoki M, Urayama T, Yamamoto I, Yamamoto I, et al. Heat sensitivity of a SARS-associated coronavirus introduced into plasma products. Vox Sanguinis. 2004;87(4):302–303.

15. Park SL, Huang YJ, Hsu WW, Hettenbach SM, Higgs S, Vanlandingham DL. Virus-specific thermostability and heat inactivation profiles of alphaviruses. J Virol Methods. 2016;234:152–155.

16. Pastorino B, Touret F, Gilles M, de Lamballerie X, Charrel RN. Evaluation of heating and chemical protocols for inactivating SARS-CoV-2. BioRxiv. doi: https://doi.org/10.1101/2020.04.11.036855

17. Pan Y, Long L, Zhang D, et al. Potential false-negative nucleic acid testing results for Severe Acute Respiratory Syndrome Coronavirus 2 from thermal inactivation of samples with low viral loads. Clin Chem. 2020;hvaa091. doi:10.1093/clinchem/hvaa091

18. Sabiiti W, Azam K, Esmeraldo E, Bhatt N, Rachow A, Gillespie SH. Heat Inactivation Renders Sputum Safe and Preserves Mycobacterium tuberculosis RNA for Downstream Molecular Tests. J Clin Microbiol. 2019;57(4).

